# Trends of per-patient healthcare cost and resource utilization of opioid use disorder among privately insured individuals in the United States from 2005-2014

**DOI:** 10.1101/2020.06.03.20121228

**Authors:** Bibo Jiang, Li Wang, Douglas Leslie

## Abstract

**Background:** Little are known about how per-patient healthcare cost and resource utilization of Opioid Use Disorder (OUD) have changed over time when governments continue to reduce availability and utilization barrier of OUD treatment.

**Objectives:** Investigate trends of per-patient healthcare cost and utilization of outpatient, inpatient and emergency department services among privately insured individuals with OUD from 2005 to 2014.

**Methods:** The MarketScan® Commercial Claims and Encounters database was used to analyze healthcare cost and resource utilization of opioid used disorder from 2005 to 2014. A matched case-control design was employed to estimate the impact of OUD on healthcare cost and service utilization over this period.

**Main findings:** Excess annual per-patient healthcare cost of OUD stayed relatively stable with an average of $14,586 between 2005 and 2014. However, excess outpatient cost increased while excess inpatient cost decreased over time. Among OUD patients, the increase of OUD related outpatient care utilization rate and average number of visits coincided with the decrease of inpatient and ED service utilization rates and average number of ED visits.

**Conclusions:** Among OUD patients, the increasing per-patient utilization of OUD related outpatient care, together with the decline in per-patient utilization of more urgent care including inpatient and emergency department care might indicate increased awareness and diagnosis of OUD and a better control of the disease among existing patients with private insurance.

## 1. Introduction

The misuse of and addiction to opioids, including prescription pain relievers, heroin, and synthetic opioids such as fentanyl, is a serious national crisis that affects public health as well as social and economic welfare in the United Sates.^1^ The nonmedical use of prescription pain relievers grew from 11.0 million to 12.5 million between 2002 and 2007. In 2010, an estimated 12.2 million people reported using pain relievers nonmedically for the first time within the past 12 months. ^3^ In recent years, sharp increase in use of illicit opioids including heroin and illicitly manufactured fentanyl has also contributed to the elevation of opioid overdose.^1^ Drug overdose deaths nearly tripled from 1999 to 2014, with over half of the drug overdose deaths in 2014 involving an opioid^1,4-6^. Opioid involved overdose death rate further increased by 15.6% from 2014 to 2015 and 27.9% from 2015 to 2016, resulting in 42,249 deaths (13.3 per 100,000 population) in 2016.^5,7^

A key driver of the overdose epidemic is underlying opioid use disorder (OUD). Consequently, expanding access to addiction-treatment services is an essential component of a comprehensive response.^8^ Like other chronic diseases such as diabetes and hypertension, addiction is generally refractory to cure, but effective treatment and functional recovery are possible.^9^

However, given the high mortality and morbidity of opioid use disorder, OUD has imposed substantial cost on payers and society and clinical burden on the healthcare system. Birnbaum et al. estimated that the total costs of prescription opioid abuse, including direct and indirect costs, reached $55.7 billion in 2007 compared with $8.6 billion in 2001.^10,11^ According to a review on studies published from 2002 to 2012 about the US economic burden of opioid abuse, when compared to a control group, mean excess healthcare cost for opioid abusers with private insurance ranged from $14.054 to $20.546 and from $5,874 to $15,183 for opioid abusers with Medicaid.^12^ In two more recent studies by Rice et al., excess annual per-patient healthcare cost of diagnosed opioid abusers was estimated as $10,627 and $11,376 using two different datasets of privately insured individuals.^13,14^

Other than researches on excess healthcare cost, there were studies focusing on clinical burdens of OUD. Studies found that in general, OUD patients utilized more healthcare services, including emergency department (ED) visits, outpatient visits and inpatient hospital stays when compared with non-OUD control subjects. With the increase of the prevalence of opioid abuse and dependence, drug treatment admissions related to prescription opioids increased more than five folds between 1998 and 2008.^15^ The rate of hospital stays involving opioid overuse among adults increased more than 150 percent between 1993 and 2012,^16^ and ED visits for opioid overdose quadrupled from 1993 to 2010.^17^

The aforementioned studies, among others, either studied the trends of the total healthcare cost or clinical burdens of OUD on the healthcare system, or estimated the excess healthcare cost of OUD by following their treatment for a short period, often 12 months after OUD diagnosis. Both types of researches did not answer the question of how per-patient healthcare cost and healthcare service utilization changed over time among diagnosed OUD patients. Firstly, the astonishing increasing trends of the total healthcare cost and clinical burdens were mainly driven by the increase of OUD prevalence, and did not reflect the change of per-patient healthcare cost for healthcare resource utilization. Secondly, because various studies on excess healthcare cost of OUD used different datasets and criteria in defining opioid abusers and selecting corresponding control groups, results from different studies are not directly comparable and therefore not very informative on how excess per-patient healthcare cost or service utilization of OUD patients changed over time.

In this paper, we used a large commercial claims database to estimate how per-patient excess healthcare cost changed from 2005 to 2014 among OUD patients with private insurance. In addition, we investigated how the utilization of different healthcare services, including outpatient, inpatient and ED services, changed over time in terms of per-patient utilization and utilization rate among diagnosed OUD patients. This is an important problem since helping OUD patients to obtain necessary treatment is a crucial aspect in battling the epidemic. In fact, both federal and state governments have put a lot effort to reduce availability and utilization barriers of OUD treatment. The availability barriers include paucity of licensed providers, capacity constraints, and stringent regulation, while utilization barriers include treatment costs and limited health insurance coverage ^9,18^.

## 2. Methods

### 2.1 Data Source and Study Sample

Data used in this study are from the MarketScan® Commercial Claims and Encounters database (MarketScan), which includes claims information from more than 130 payers, and describes the health care service use and expenditures for tens of millions (varying from year to year) of covered employees and family members per year. The database is divided into subsections, including inpatient claims, outpatient claims, outpatient prescription drug claims, and enrollment information. Claims data in each of the subsections contain a unique encrypted patient identifier and information on patient age, sex, geographic location (e.g., state), etc.

The MarketScan dataset used in this study covers the period from January 2005 through December 2014 for all 50 states. The demographic characteristics of the study sample are presented in Table 1. Table 1 shows that the sample does not contain any individuals aged 65 and older, therefore, the results presented in this paper are for a privately insured non-elderly (aged 64 and younger) cohort. In addition, Table 1 shows that the demographic characteristics of included individuals remained relatively stable over this period. Institutional review board approval was obtained prior to implementation of the study.

**Table 1:**
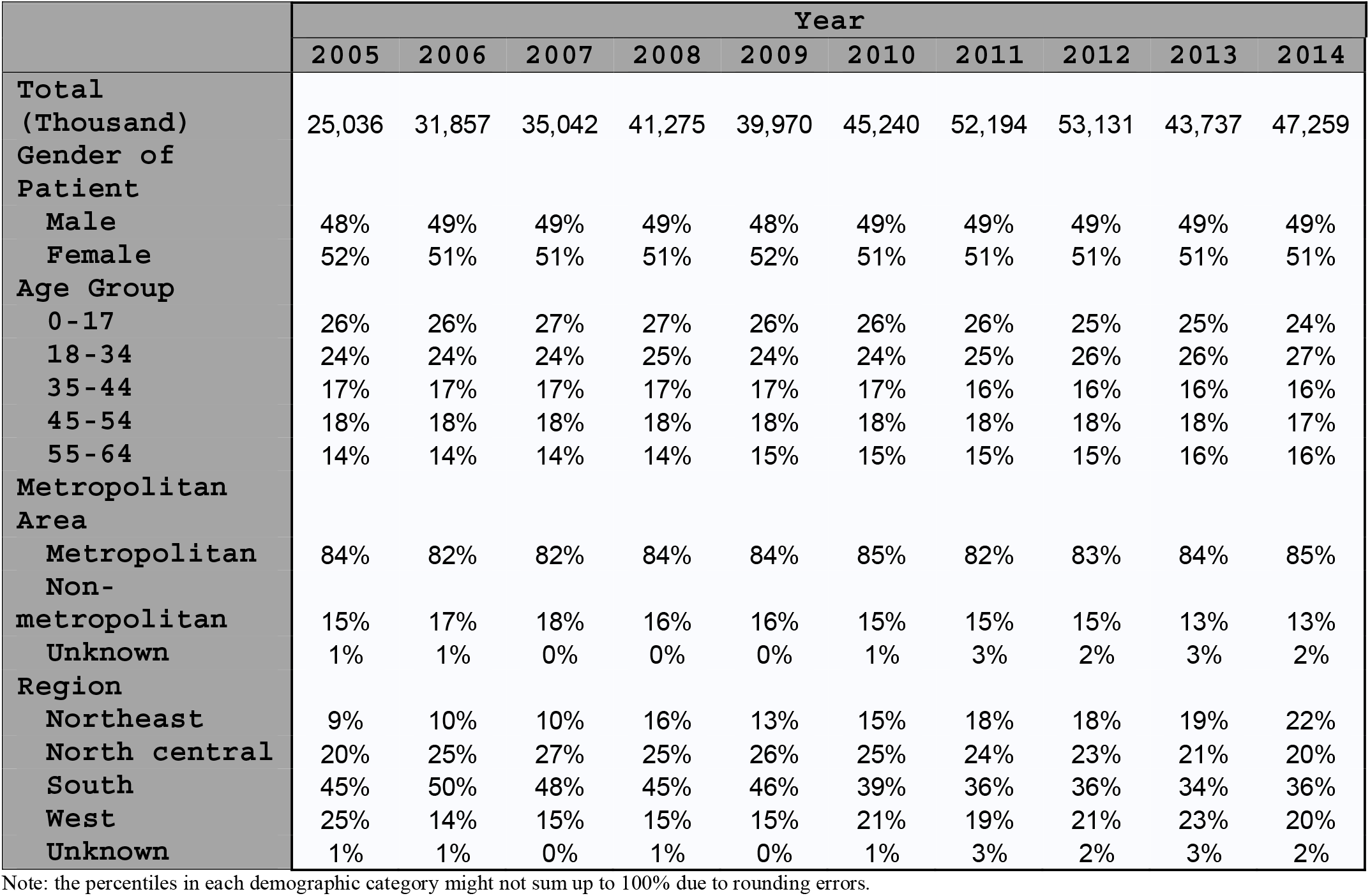
Demographic statistics of enrollments in the MarketScan database

### 2.2 Study Measures and Analysis

Opioid use disorder is a diagnosis introduced in the fifth edition of the Diagnostic and Statistical Manual of Mental Disorders (DSM-5). It combines two disorders from the previous edition of the Diagnostic and Statistical Manual, the DSM-IV-TR, known as Opioid Dependence and Opioid Abuse, and incorporates a wide range of illicit and prescription drugs from the opioid class. Following previous studies^19,20^, we identified OUD patients for each year as those having at least one inpatient or outpatient claim with a primary ICD-9 diagnosis code 304.0x (opioid type dependence), 304.7x (combinations of opioid type drug with any other drug dependence) and 305.5x (nondependent opioid abuse) in that year. Table 2 presented the number and demographic characteristics of identified OUD patients for each year. Due to different increasing rates of OUD prevalence in different subgroups, such as age group, gender and location etc., the demographic characteristics of identified OUD patients did change over time.

**Table 2:** Demographic Characteristics of OUD Patients (2005 - 2014)

|  | Year |  |  |  |  |  |  |  |  |  |
| --- | --- | --- | --- | --- | --- | --- | --- | --- | --- | --- |
|  | 2005 | 2006 | 2007 | 2008 | 2009 | 2010 | 2011 | 2012 | 2013 | 2014 |
| **Total (N)** | 9884 | 14026 | 17737 | 27835 | 32076 | 44053 | 67839 | 81422 | 76237 | 95160 |
| **Gender of Patient** |  |  |  |  |  |  |  |  |  |  |
| **Male** | 57.5% | 58.4% | 58.7% | 60.0% | 59.7% | 60.8% | 62.2% | 62.7% | 62.4% | 61.9% |
| **Female** | 42.5% | 41.6% | 41.3% | 40.0% | 40.3% | 39.2% | 37.8% | 37.3% | 37.6% | 38.1% |
| **Age Group** |  |  |  |  |  |  |  |  |  |  |
| **0-17** | 4.1% | 4.3% | 3.7% | 3.2% | 3.0% | 2.7% | 2.2% | 1.8% | 1.5% | 1.0% |
| **18-34** | 37.3% | 39.7% | 42.2% | 44.1% | 46.5% | 48.7% | 55.4% | 55.6% | 54.5% | 52.2% |
| **35-44** | 23.2% | 22.5% | 22.0% | 21.5% | 20.2% | 20.5% | 18.4% | 18.4% | 18.5% | 19.5% |
| **45-54** | 26.5% | 24.1% | 23.0% | 21.9% | 20.6% | 18.9% | 16.0% | 15.6% | 15.4% | 15.8% |
| **55-64** | 8.9% | 9.3% | 9.1% | 9.4% | 9.8% | 9.3% | 8.1% | 8.6% | 10.2% | 11.4% |
| **Metropolitan Area** |  |  |  |  |  |  |  |  |  |  |
| **Metropolitan** | 87.3% | 86.1% | 86.2% | 87.5% | 86.4% | 83.7% | 81.6% | 82.9% | 84.0% | 84.1% |
| **Non-metropolitan** | 11.5% | 13.1% | 13.5% | 12.2% | 13.4% | 15.2% | 15.3% | 15.2% | 13.7% | 13.8% |
| **Unknown** | 1.2% | 0.7% | 0.3% | 0.3% | 0.2% | 1.1% | 3.1% | 1.9% | 2.3% | 2.0% |
| **Region** |  |  |  |  |  |  |  |  |  |  |
| **Northeast** | 12.0% | 15.0% | 14.3% | 23.2% | 16.4% | 22.0% | 27.0% | 28.4% | 29.2% | 31.4% |
| **North central** | 24.8% | 28.2% | 27.4% | 22.6% | 23.4% | 22.7% | 21.7% | 19.5% | 18.5% | 17.4% |
| **South** | 43.5% | 42.3% | 42.6% | 38.7% | 43.1% | 36.3% | 32.6% | 33.3% | 31.4% | 34.1% |
| **West** | 18.4% | 13.8% | 15.3% | 15.2% | 16.9% | 17.9% | 15.6% | 16.9% | 18.5% | 15.0% |

Note: the percentiles in each demographic category might not sum up to 100% due to rounding errors.

To accommodate the change of demographic characteristics of OUD patients over time and study the change of healthcare cost and resource utilization of OUD patients, we used a matched case-control study approach. Each year, a non-OUD control group was randomly selected from the enrollment database based on 1:1 exact matching of gender, age, state and metropolitan/nonmetropolitan. Therefore, each year the distribution of the non-OUD control group in terms of these variables was exactly the same as that of the OUD group.

## 3. Results

### 3.1 Trends of Healthcare cost of OUD Patients

Figure 1 depicts the trends of healthcare cost, including impatient, outpatient and drug costs, for OUD and non-OUD patient. The costs were adjusted using the CPI for medical goods and services (base year: 2014) reported by Bureau of Labor Statistics. The left panel shows the trend of total healthcare cost of OUD patients (per million enrollments)^2^. The total healthcare cost (summation of all inpatient, outpatient and drug costs) for OUD patients increased from 8.0 million dollars in 2005 to 42.1 million dollars in 2014, a little more than 5 folds. Among which, OUD related costs (summation of healthcare cost from inpatient and outpatient claims with OUD diagnosis as principle diagnosis and OUD related drug^3^ cost) increased from 1.5 million to 11.3 million dollars during this period, more than 7 folds. OUD prevalence rate^4^ increased from 0.04% to 0.20% during this period.

**Figure 1:**
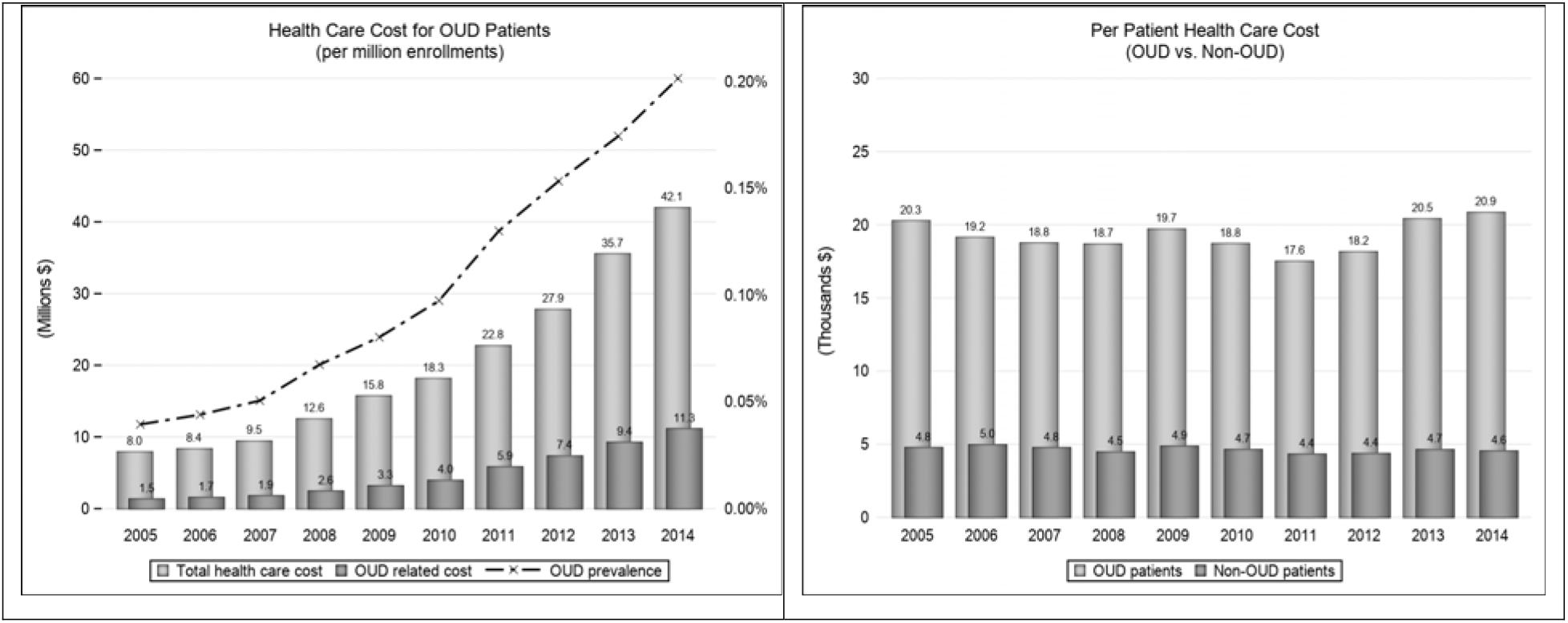
Medical costs of OUD patients vs. non-OUD patients (2005 – 2014)

The right panel of Figure 1 depicts per-patient healthcare cost for OUD patients during this period. As a comparison, we also plotted per-patient healthcare cost for the control group alongside. As we can see, both series stayed relatively stable during this period, with average per patient health care costs for OUD patients being $19,265 compared with $4,679 for the control group. The average excess annual per-patient healthcare cost was $14,586.

To further investigate different components of healthcare cost for OUD patients, we plotted the trends of per-patient inpatient, outpatient and drug costs of OUD patients in Figure 2. Again, the corresponding cost components of non-OUD patients were plotted alongside for comparison. For OUD patients, per-patient inpatient cost trended downward considerably from $8,017 (39.4% of the total costs) to $5,725 (27.4% of the total costs) as per-patient outpatient cost increased considerably from $8,927 (43.9% of the total costs) to $12,027 (57.6% of the total costs) from 2005 to 2014. The per-patient drug costs stayed stable, with average per-patient drug cost being $3,083 (16.0% of the total costs). For the non-OUD group, there were no evident trends on the change of the three cost components. The annual excess per-patient inpatient cost decreased from $6,816 in 2005 to $4763 in 2014 as excess per-patient outpatient cost increased from $6,352 to $9,479. The excess per-patient drug cost varied around $2,105.

**Figure 2:**
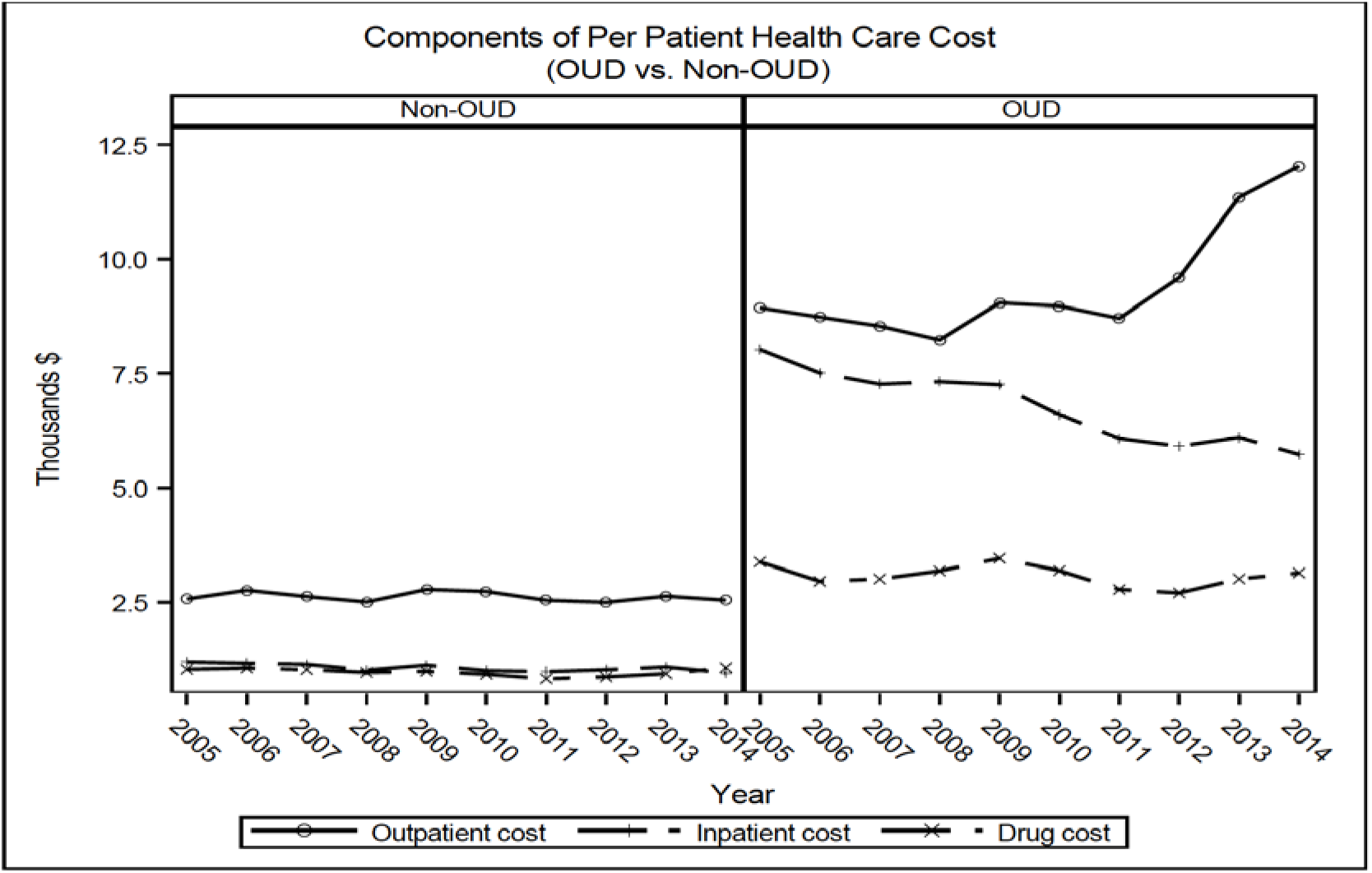
Average per-patient inpatient care, outpatient care and drug costs of OUD patients vs. non-OUD patients (2005 – 2014)

To better understand the change of outpatient and inpatient costs for OUD patients, we calculated the costs based on two categories: OUD related costs (from claims with OUD disagnosis as prince diagnosis) and other medical costs. We can see from Figure 3 that per-patient inpatient cost of both categories trended downward over this period. The per-patient OUD related outpatient cost did not change much from 2005 to 2008, and increased considerably afterwards. The not-OUD related outpatient cost fluctured before 2011, and experienced a rapid increase afterwards.

### 3.2 Trends of Healthcare Service Utilization of OUD Patients

In this section, we presented the trends of healthcare service utilization of OUD patients. In particular, we investigated inpatient, ED and outpatient care utilization of OUD patients, compared with the non-OUD group.

**Figure 3:**
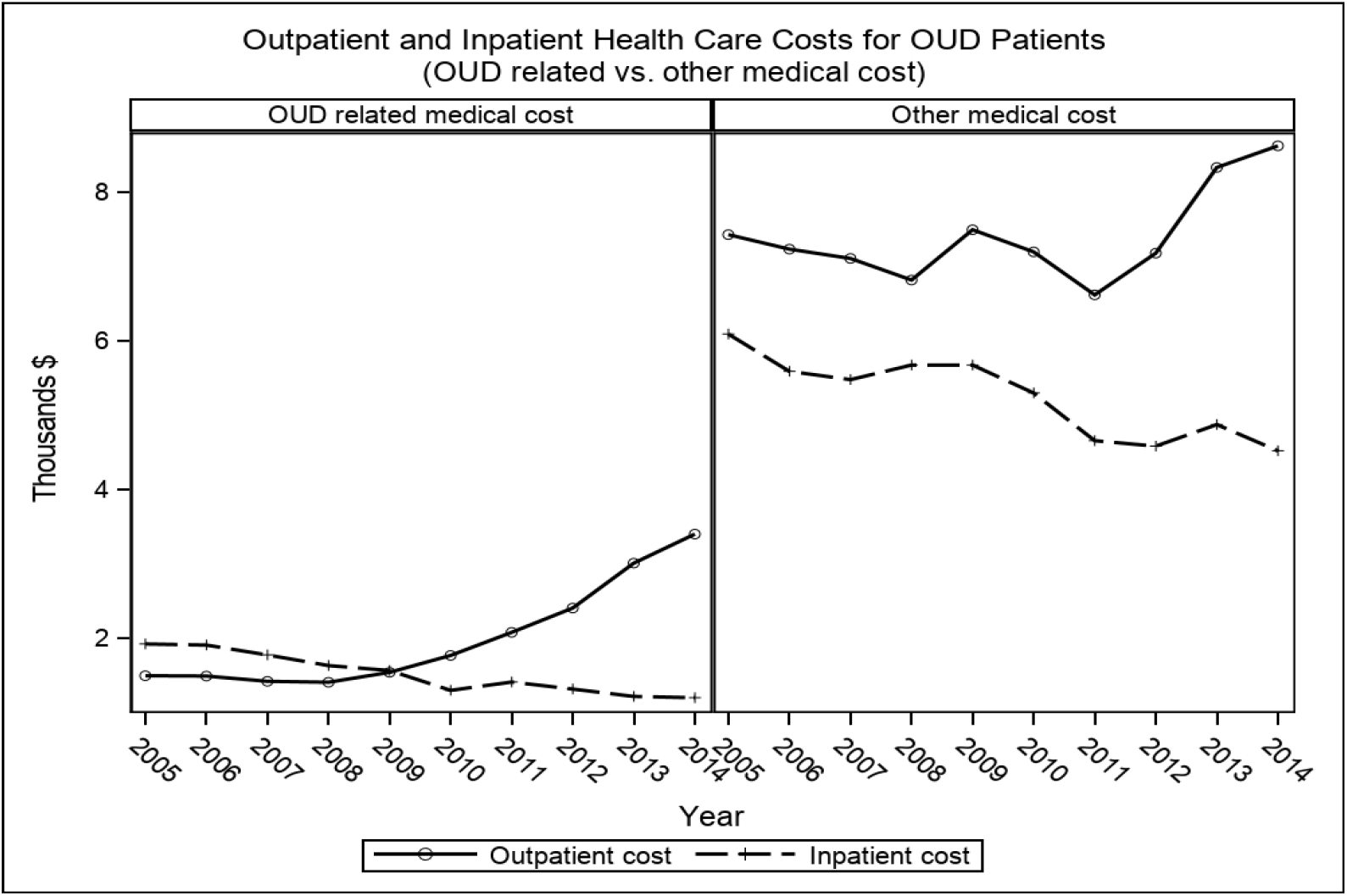
Per-patient Inpatient and Outpatient Costs of OUD patients

#### 3.2.1 Inpatient Care and Emergency Department Visits

The left panel of Figure 4 shows the average hospitalization days of OUD patients (total hospitalization days / total number of OUD patients who were hospitalized at least once during that year) and utillization rate of inpatient service (number of OUD patients who were hospitalized at least once / total number of OUD patients identified in the same year). As a comparison, we also presented the average hospitalization days and inpatient service utilization rate for the control group on the left panel of Figure 4. Similarly, on the right panel of Figure 4, we reported average number of ED visits (total ED visits / total number of OUD patients who had at least one ED visit during that year) and ED utilization rate (number of OUD patients who had at least one ED visit / total number of OUD patients identified in the same year).

**Figure 4:**
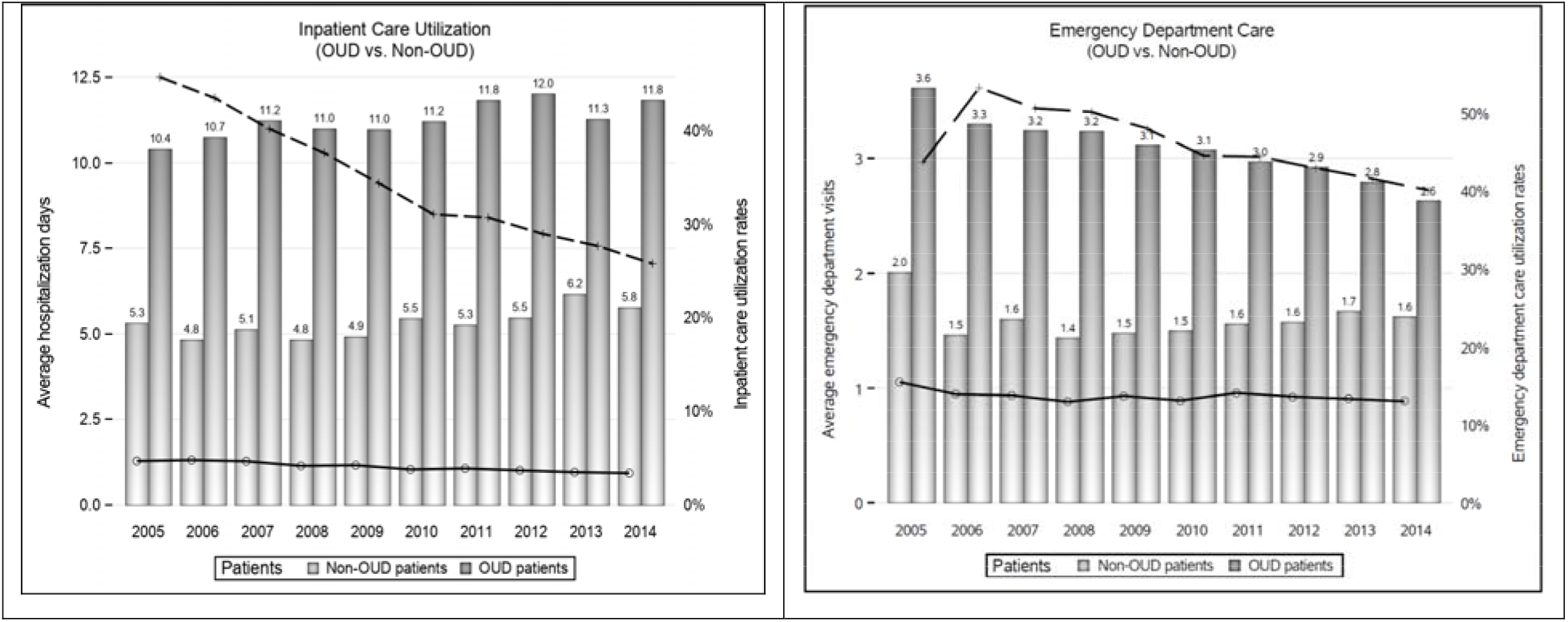
Utilization of inpatient care and ED service of OUD patients vs. non-OUD patients

From the left panel we can see that inpatient care utilization rate of OUD patients decreased dramatically during this period from 45.7% in 2005 to 25.8% in 2014, but the average hospitalization days trended upward slightly, from 10.4 days in 2005 to 11.8 days in 2014 with some flucturation between the two years. During the same period, there was a slight decrease of inpatient care utilization rate for non-OUD patients from 4.7% in 2005 to 3.4% in 2014. The average hospitalization length of non-OUD patients also experienced a slight increase from 5.3 days in 2005 to 5.7 days in 2014 with some flucturation between the two years. The utilization rate of inpatient care droped 19.9% (45.7% of the level in 2005) for OUD patients, compared with 1.3% (27.7% of the level in 2005) for non-OUD individuals.

The right panel of Figure 4 shows the trends of ED utilization of OUD patients from 2005 to 2014 compared with non-OUD individuals. There was a decreasing in utilization rate of ED service (except for 2005^5^), as well as number of per-patient ED visit for OUD patients who utilized ED service in that year. The ED service utilization rate decreased from 53.2% in 2006 (43.6% in 2005) to 40.0% in 2014, and per-patient ED visits decreased from 3.3 in 2006 (3.6 in 2005) to 2.6 in 2014. For non-OUD individuals, the utilization rate of ED service did not show any systematic change and it flucturated around its average 13.7%. However, per-patient ED visits did increse slightly in general (except for 2005) from 1.5 in 2006 (2.0 in 2005) to 1.6 in 2014 with some flucturation between the two years. To summarize, although OUD patients utilized much more ED service than non-OUD individuals, the decrease of the utilization both in terms of utilization rate and per-patient ED visits, was magnificent among OUD patients. There was no sign of decreasing utilization rate of ED service among non-OUD patients and the number of per-patient ED visit even experienced a slight increase during this period.

#### 3.2.2. Outpatient care

In this section, we investigated outpatient care utilization of OUD patients comppared with non-OUD individuals. Outpatient care utilization rate (number of OUD patients who had at least one outpatient claim / total number of OUD patients identified in that year) and number of per-patient outpatient visits (total number of outpatient visits / number of OUD patients who had at least one outpatient claim during that year) were presented on the left panel of Figure 5. The corresponding measures for non-OUD individuals were presented alongside. From the left panel of Figure 5, we can see that the outpatient care utilization rate among OUD patients was above 99% throughout the period. This is not superising since each year we defined OUD patients as those who had at least one inpatient or outpatient claim with an OUD diagnosis as principle diagnosis. It is rare that OUD patients identified through inpatient claims never had any outpatient claims. The outpatient care utilization rate for non-OUD individuals was about 75%.

**Figure 5:**
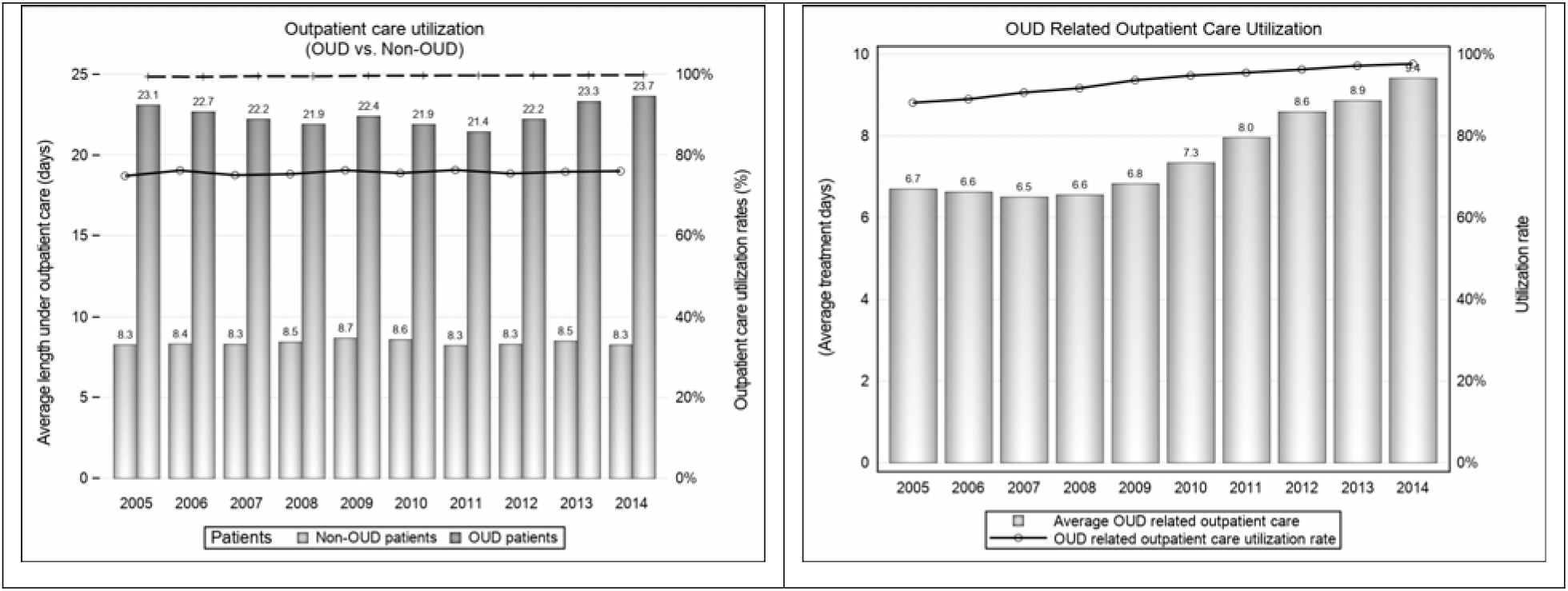
Out patient service utilization of OUD patients vs. non-OUD patients

The number of per-patient outpatient visits for OUD patients exhisibited a pattern of first decreasing and then increasing with an average number of visits being 22.5, compared with 8.42 for non-OUD individuals.

To further investigate the utilization of outpatient service among OUD patients, we looked into utilization of OUD related outpatient care in particular. We plotted the OUD related outpatient utilization rate and number of per-patient visits on the right panel of Figure 5. The utilization rate increased from 88.1% in 2005 to 97.6% in 2014 which implies that some OUD patients identified from inpatient claims never had OUD related treatment through outpatient care in that year although this percentage decreased during this period. The number of per-patient visits stayed stable from 2005 to 2008 and increased significantly since 2009, reaching 9.4 in 2014 compared with 6.7 in 2005. The number of per-patient visits increased 2.7, 40.6% of the level in 2005.

## 4. Discussions

The total healthcare cost for OUD patients reached 42.1 million dollars in 2014, a little more than 5 folds of 8.0 million dollars in 2005. During this sample period, OUD prevalence rate increased from 0.04% to 0.20%, with an increasing rate similar to that of the total healthcare cost. This implies that the increase of the total healthcare cost of OUD was mainly driven by the increase of OUD prevalence, which further explains why per-patient healthcare cost for OUD patients did not change much during this period. The average excess annual per-patient healthcare cost varied around its average $14,586.

It is worth noting that OUD related healthcare cost increased at a faster rate than the overall healthcare cost for OUD patients. As a result, the percentage of OUD related cost among overall healthcare cost increased from 18.8% in 2005 to 26.8% in 2014. This indicates that more and more medical resources were used in the treatment of the disease itself. However, even in 2014, a major portion (73.2%) of the healthcare cost of OUD patients were claimed from treatment of other health problems. Therefore, reducing ricks of contaminating serious OUD morbidities, such as HIV, is crucial to control healthcare cost of OUD patients. A better control of the disease itself may potentially reduce the risk of developing mobidities.

Although per-patient healthcare cost of OUD did not change much from 2005 to 2014, the distribution of the costs among outpatient, inpatient and drug utilization changed over time.

The per-patient outpatient cost increased from $8,927 (43.9% of the total costs) to $12,027 (57.6% of the total costs) and the per-patient inpatient cost decreased from $8,017 (39.4% of the total costs) to $5,725 (27.4% of the total costs). This is consitent with the findings on utilizations of inpatient and outpatient services among OUD patients.

Inpatient care utilization rate of OUD patients decreased dramatically during this period, from 45.7% in 2005 to 25.8% in 2014 although the average hospitalization days among those who used inpatient services increased slightly from 10.4 days in 2005 to 11.8 days in 2014. The big drop of the inpatient care utilization rate is likely the reason for the significant decrease of per-patient inpatient cost of OUD patients. During the same period, there was a slight decrease of inpatient care utilization rate for the non-OUD group, from 4.7% in 2005 to 3.4% in 2014 and similarly the average hospitalization lenth slighly increased from 5.3 days in 2005 to 5.7 days in 2014. For both groups, the decrease on inpatient care utilization rates and increase on average hospitalization days among those who used inpatient services might partially due to growing efforts on reducing unnecessary hospitalizations, greater use of chronic disease management programs, and a shift toward outpatient treatment. Those who did not need to be hospitalized were referred to outpatient service which made the inpatient utilization rate lower. Those being hospitalized were likely to be more illed and therefore average hospitalization days increased.

However, this general trend toward utilizing more outpatient care might not be sufficient in explaining the dramatic drop of inpatient care utilization rate from OUD patients. The utilization rate of inpatient care dropped 19.9% (45.7% of the level in 2005) for OUD patients, much more significant than 1.3% (27.7% of the level in 2005) for non-OUD individuals. Of course, the decline could also be caused by OUD treatment preference switching from inpatient to outpatient, for example, from inpatient rehab to outpatient rehab. Although we did not find evidence for such a treatment preference shift, it could still exist. However, the fact that per-patient inpatient cost for both treating OUD and other health problems declined (Figure 3) significantly during this period indicated that OUD treatment preference shift could not fully explain the slidedown even if such a shift did exis.

Although the decrease of inpatient service utilization might be due to a shift toward outpatient treatment, the evident decrease of ED utilization among OUD patients (both in terms of utilization rate and average number of visits among those who utilized ED service) likely indicated a better control of health condictions among OUD patients since ED service is designed to handle acute, and even fatal health conditions. It is less likely to be affected by shift of treatment preference. Moreover, this pattern did not appear in the non-OUD control group.

Contrary to the decrease of inpatient and ED care utilization, there was an increase in both OUD related outpatient care utilization rate and average number of OUD related outpatient visits among those who reveived OUD treatment from outpatient care. The increase of average number of visits from 2009 (Figure 5) coincided with the encatment of the Paul Wellstone and Pete Domenici Mental Health Parity and Addiction Equity Act of 2008 (MHPAEA). It is a federal law that generally prevents group health plans and health insurance issuers that provide mental health or substance use disorder (MH/SUD) benefits from imposing less favorable benefit limitations on those benefits than on medical/surgical benefits.^22^ MHPAEA, together with Patient Protection and Affordable Care Act reduced availability barrier of OUD treatment and likely encouraged OUD patients to seek OUD related treatment more proactively. On the one hand, this could help reduce the percentage of undiagnosed OUD patients in privately insured population. Kirson et.al found that the ratio of undiagnosed to diagnosed abuse declined considerably from 2006 to 2011 among commercially insured individuals, and the ratio was 2:1 in 2011. ^23^ Another study found a similar trend of increasing rates of diagnosed opioid abuse among commercially insured individuals continuing into 2012. ^14^ When more new OUD patients with relatively good health condition were diagnosed and joined the cohort, the utilization rate of inpatient care and ED service among OUD patients were likely to decrease. On the other hand, more proactively seeking OUD treatment could also help existing OUD patients to better manage the disease, reduce the risk of developing morbidities, and avoid using ED or inpatient services.

Although not of the focuse of this paper, it is worth noting that as more outpatient services and less inpatient and ED services were used, the indirect costs of the disease, such as workday loss from OUD patients and their family members, were likely to decrease, although excess per-patient healthcare cost of OUD patients did not decrease (nor increase) over this period.

## 5. Limitations

The findings in this research are subject to the following limitations. First, the study was based on claims data from MarketScan, which is a large convenient sample but not nationally representative. The trends of per-patient healthcare cost and service utilization presented in this paper might not reflect the corresponding trends in privately insured population of U.S. More researches to study per-patient healthcare cost and service utilization by using different samples would be helpful.

In addition, this study did not specially examine the factors that caused the change of per-patient healthcare cost and service utilizations, instead, focused on identifying trends on per-patient healthcare cost and service utilization among OUD patient with private insurance. The finding that increasing utilization rate of OUD-related outpatient service coincided with decreasing utilization rate on ED and inpatient service might indicate a better manage of the disease among OUD patients. However, other factors, such as changing demographic characteristics of OUD patients over time, might also contribute to the decrease of inpatient and ED utilization rates, although we did not see such pattern in the control group. Further investigation on this issue will be helpful to fully understand the driving horses of the trends.

Finally, like all studies based on claims data, our study was not able to capture those OUD patients who never sought for treatment. This might render overestimation of the utilization rates for inpatient, ED and outpatient services among OUD patients with private insurance.

## 6. Conclusions

The increase of the total healthcare cost of OUD patients with private insurance from 2005 to 2016 was mainly driven by the increase of OUD prevalence. Excess annual per-patient healthcare cost stayed relatively stable over this period. Among OUD patients, the increasing per-patient utilization of OUD related outpatient care, together with the decline in per-patient utilization of more urgent care including inpatient and ED care, might indicate increased awareness and diagnosis of OUD and a better control of the disease among existing patients with private insurance. Efforts on continuing reducing availability and utilization barrier of OUD treatment is crucial in combating the opioid epidemic.

## Data Availability

Data used in the study come from the MarketScan Commercial Claims and Encounters database. Although we are not allowed to share the data, the data are publicly available with purchase.

## Footnotes

2 To adjust for cross-year variation of the number of enrolled individuals in the database, the reported health care costs were based on 1 million enrollments.

3 OUD related drugs include methadone, buprenorphine and naltrexone. Although these drugs could be prescribed for treatment of other addictions, such as alcohol addiction, we still assumed that these drugs prescribed for OUD patients were used for OUD treatment since no information was available in the dataset to identify prescribing purpose.

4 For each year, OUD prevalence is defined as the total number of OUD patients divided by the total number of enrollments.

5 ED service utilization in 2005 for both OUD and non-OUD groups behaved differently from other years in the sample period. We failed to find any good explanation for that. One possible reason could be that the sample size in 2005 was much smaller than in other years, indicating more payers were included in the dataset in later years. Because ED visits are rare cases, the statistics based on ED visits could be more affected by the change of sample than statistics on inpatient and outpatient service utilization. Because of this reason, we presented the trends using results from 2006 and 2014, with results from 2005 included in parentheses.

